# Sociodemographic and health correlates of reimbursement authorizations for cannabis for medical purposes in Canadian veterans: A cross-sectional study linking the *Life After Services Studies 2019* and *Health Administrative Databases*

**DOI:** 10.64898/2026.06.10.26355368

**Authors:** Tetyana Kendzerska, Julián Reyes, Noah Poirier, Alain Poirier, Alex Cull, Anthony Murkar, Mouaz Saymeh, Stephanie Belanger, Monnica Williams, Jakov Shlik, Rakesh Jetly, Rebecca Robillard

## Abstract

**Background:** Evidence on factors associated with cannabis for medical purposes (CMP) authorizations among Veterans Affairs Canada (VAC) clients remains limited and inconsistent, particularly concerning mental health and posttraumatic stress disorder (PTSD), a leading indication for use. We investigated demographic, clinical and service characteristics associated with VAC authorizations for CMP reimbursement.

**Method:** We linked VAC administrative CMP program data with responses from the 2019 Life After Services Studies cross-sectional survey of Regular Force veterans released between 1998 and 2018. Multivariable logistic regressions examined associations between CMP reimbursement (yes/no) and demographic, clinical and well-being factors, with analyses stratified by PTSD status.

**Results:** Among 1,289 respondents (weighted n=33,131), 18.4% were authorized for CMP reimbursement. Younger age (<40 vs. ≥60 years: OR 4.78, 95% CI: 2.24–10.21), unemployment with inability to work vs. employed (OR 3.10, 95% CI: 1.78–5.40), land service vs. air (OR 2.07, 95% CI: 1.22–3.50), PTSD (OR 2.81, 95% CI: 1.69–4.66), anxiety (OR 2.32, 95% CI: 1.45–3.70), and severe pain vs. no pain (OR 3.61, 95% CI: 1.97–6.60) were independently associated with authorization. Unemployment and severe pain were consistent correlates across PTSD strata. Among those without PTSD, younger age, multiple physical conditions, and frequent mental health visits were significant; among those with PTSD, shorter service, witnessing destruction, and suicidal ideation were additional factors.

**Conclusions:** CMP authorization patterns among Canadian veterans reflect the intersection of mental health, pain, and functional impairment, with variation by PTSD status. These findings underscore the need for longitudinal research on CMP mechanisms, effectiveness and safety.

## Introduction

In recent years, Veterans Affairs Canada (VAC) has seen a substantial increase in authorizations for reimbursement for cannabis for medical purposes (CMP) [1,2]. At the same time, current evidence on factors associated with CMP prescription remains limited and heterogeneous, especially in the context of mental health and posttraumatic stress disorder (PTSD), one of the lead motives of use for medical CMP in veterans [3–5]. Understanding these factors is critical because medical cannabis is increasingly prescribed for a wide range of physical and mental health conditions, often without robust supporting evidence, raising concerns about optimal therapeutic use, patient safety, and the risk of adverse effects or drug interactions [6–11]. This is even more important for the veteran population because of the high burden of other CMP-related conditions in addition to PTSD, like chronic pain, sleep issues, and psychiatric comorbidities, the risk of substance use disorders, and the need for safe, effective, and evidence-based management strategies in this vulnerable population with complex health needs [1,3,12]. Improved understanding of factors associated with CMP prescription and authorization can inform prescriber education, regulatory policy, and the development of evidence-based guidelines to ensure that medical cannabis is used appropriately and safely.

In the general population, factors associated with CMP prescribing include demographic characteristics (younger age; male sex for THC-dominant products and older age or female sex for CBD-dominant products), clinical indications (chronic pain, cancer-related symptoms, anxiety, sleep disorders, PTSD, and depression), and concurrent medication use (e.g., opioids, antipsychotics) [6,7,9]. Socioeconomic status, access to alternative treatments, and regional regulatory frameworks may also shape prescribing practices for CMP [13,14]. Studies on factors associated with the CMP prescription in veterans demonstrated both overlap and important differences compared to the general population. In veterans, younger age, male sex, lower income, hazardous alcohol use, chronic pain, and mental health conditions, such as PTSD, depression, or anxiety, have been shown to be associated with higher rates of medical cannabis use and prescription [15–18]. Compared to the general population, veterans are also more likely to use cannabis as a substitute for prescription medications, particularly in the context of polypharmacy and opioid use [3]. However, many existing studies rely on self-reported data, lack validated assessment tools, do not differentiate medical from recreational cannabis use, or have limited adjustment for confounding factors and exclude marginalized populations [6,7,9].

Thus, to address the existing knowledge gap, this study aims to help understand how specific characteristics interact with the CMP prescription among veterans by investigating the associations between receiving authorizations for VAC reimbursement for CMP and clinical, demographic, and service characteristics, using administrative information and the 2019 Life After Services Studies (LASS) survey.

## Materials and Methods

### Study Design and Data Sources

This study utilized linked administrative data on VAC authorization for CMP reimbursement and responses from the 2019 LASS, a cross-sectional survey that collected information on veterans released from the Regular Force between 1998 and 2018 [19]. VAC may provide reimbursement for CMP when it is both authorized by a healthcare provider and obtained in compliance with Health Canada’s Cannabis Regulations, in line with VAC’s policy on CMP reimbursement. LASS 2019 was representative of 79,997 veterans, with a response rate of 72% and a response share of 92% [19]. This survey was stratified into three groups by rank at release: officers, senior non-commissioned members (Sr NCM), and junior non-commissioned members (Jr NCM). All identifiers were removed after the data linkage procedure. Figure S1 shows the collection of data sources used in the present analysis.

### Ethics approval

Participants who consented to linkage with VAC administrative data were included in this study. Data collection and access procedures for the LASS 2019 survey were reviewed and approved by Statistics Canada policy committees, which serve a role analogous to a research ethics board [20]. Ethics approval for this project was granted by the University of Ottawa Research Ethics Board (H-02-22-7491). All procedures complied with the ethical standards of relevant national and institutional committees on human experimentation and with the 1975 Helsinki Declaration, as revised in 2013. Written informed consent was obtained from all survey participants.

### Variable Selection and Definitions

The cannabis authorization variable was dichotomized (yes versus no) based on whether or not a veteran had an active authorization for CMP as of March 2019. Variables were selected through a literature review and experts’ opinions (obtained via personal communication and internal discussions among coinvestigators) from an initial list of 57 variables (see details in Table 1 and Table S1).

**Table 1.**
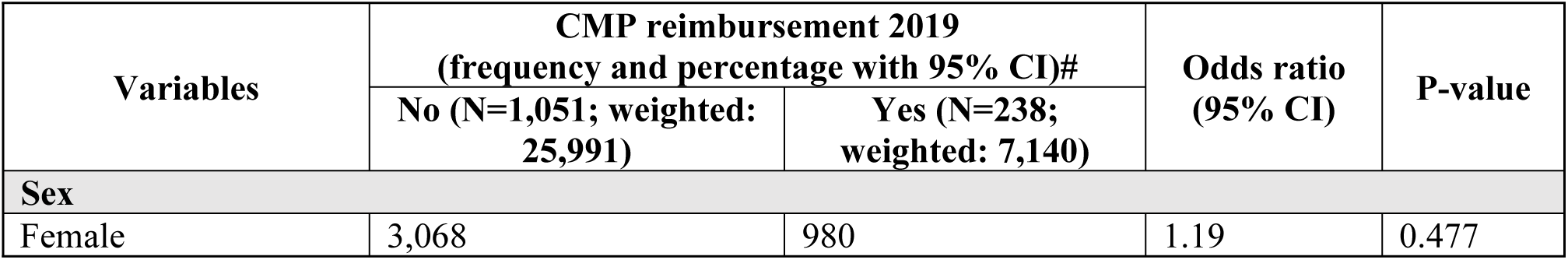
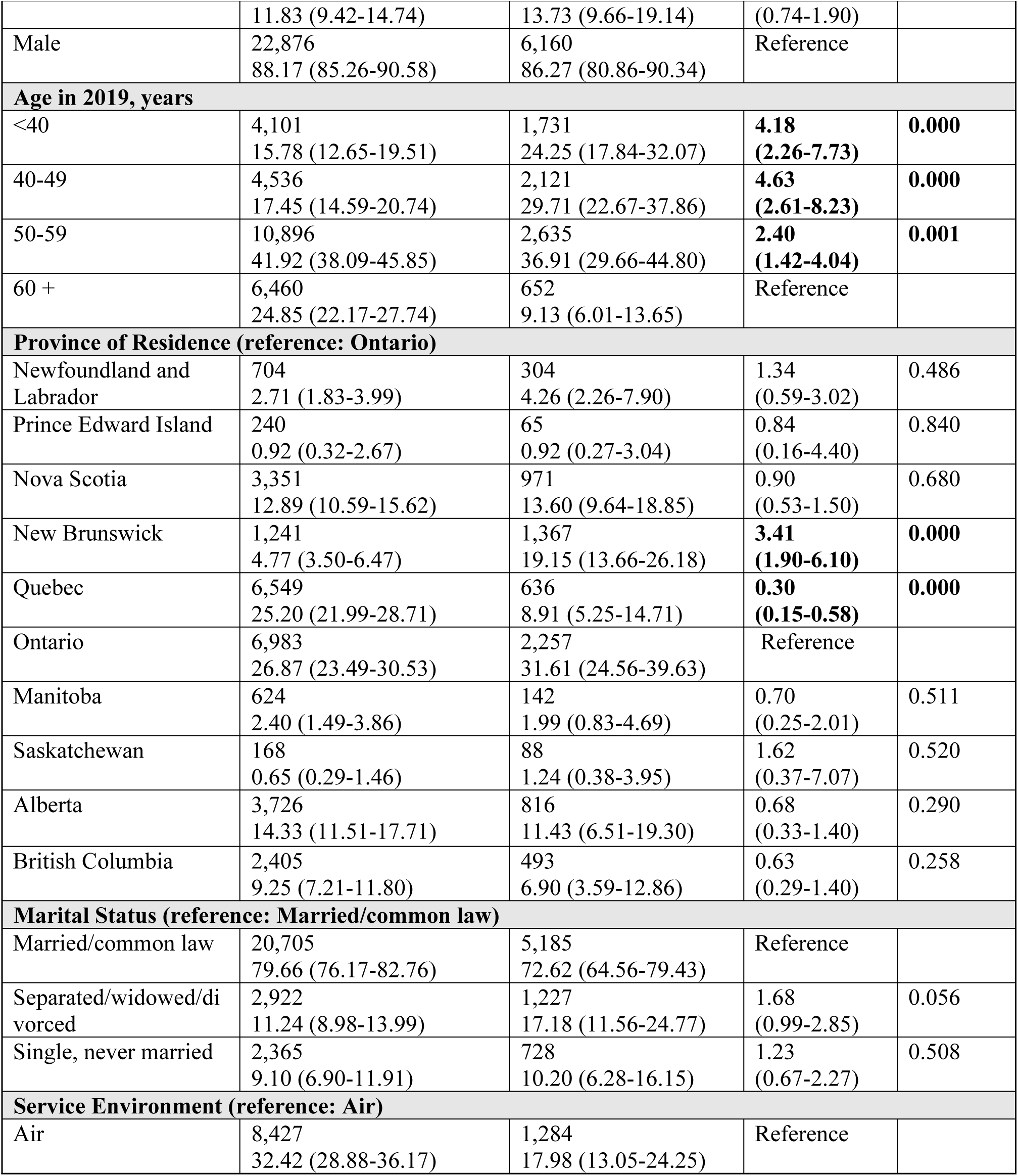
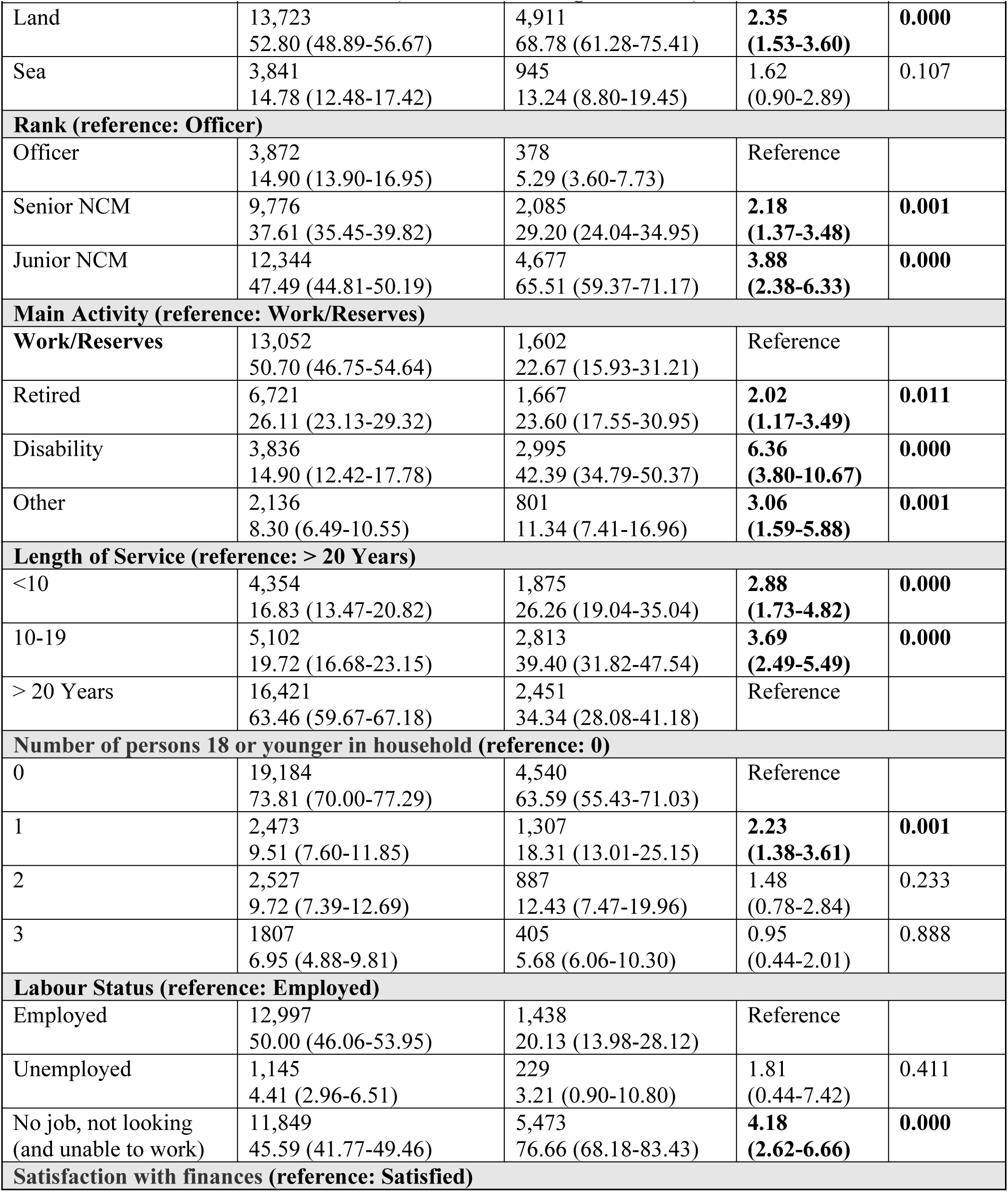
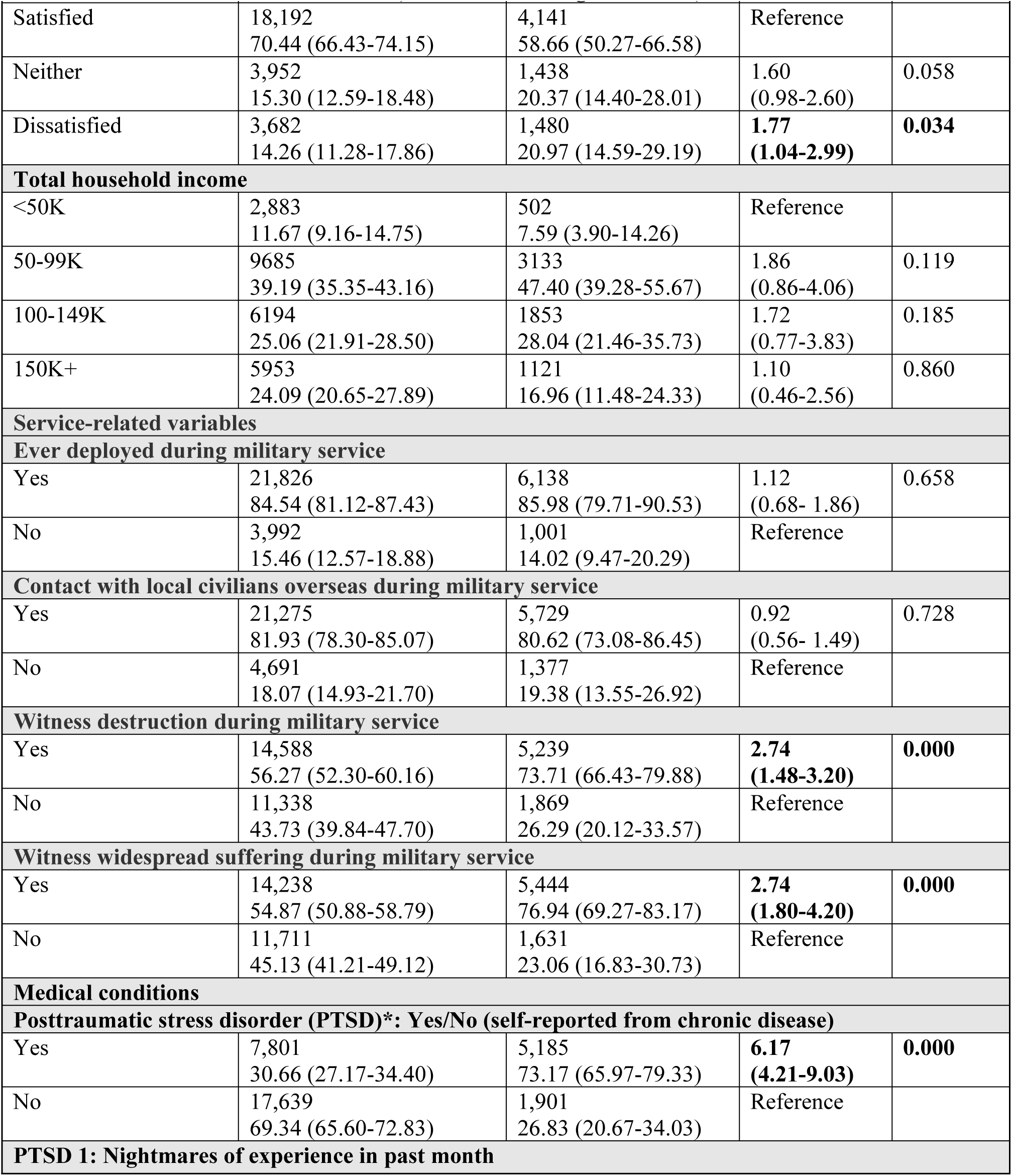
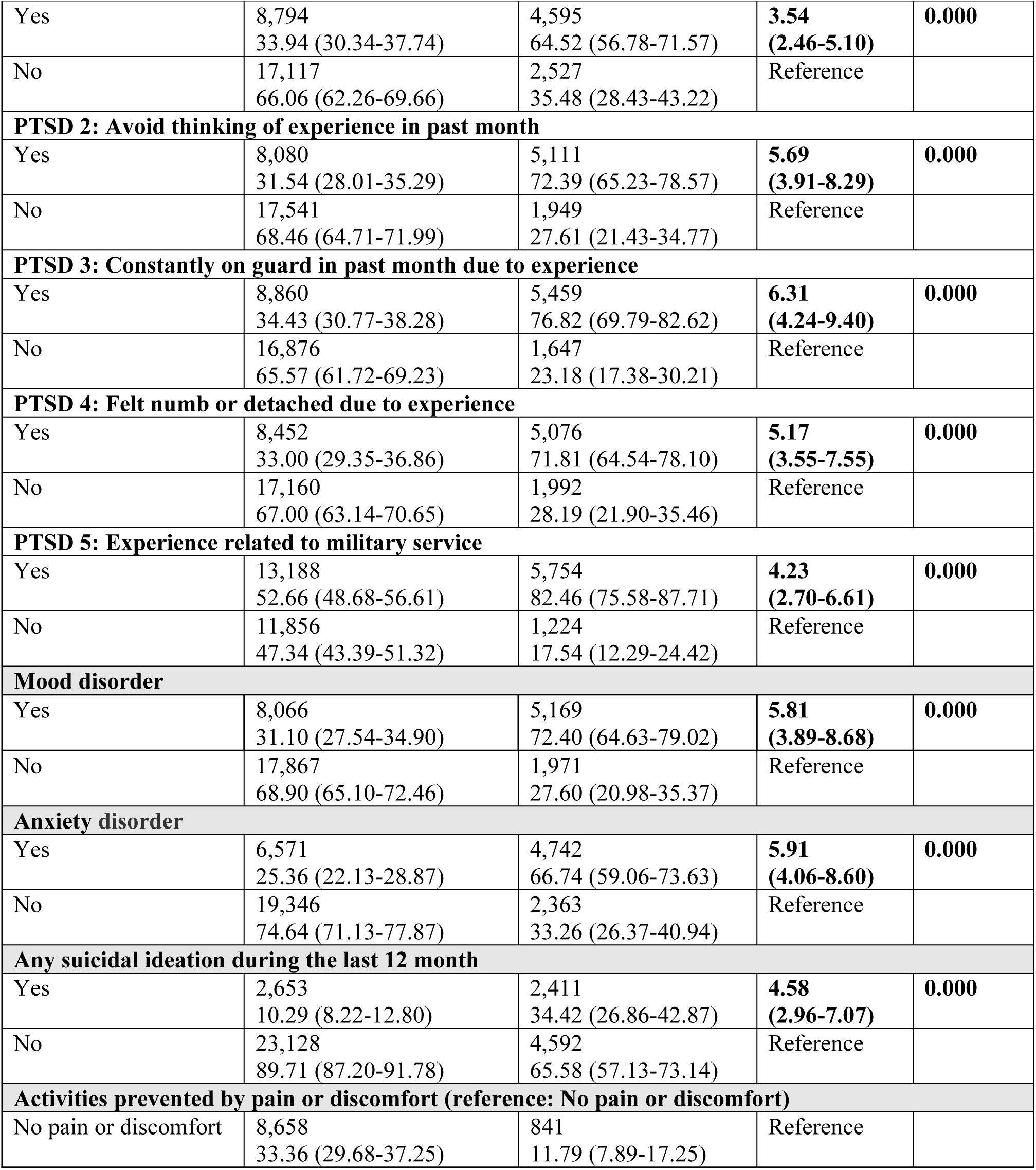
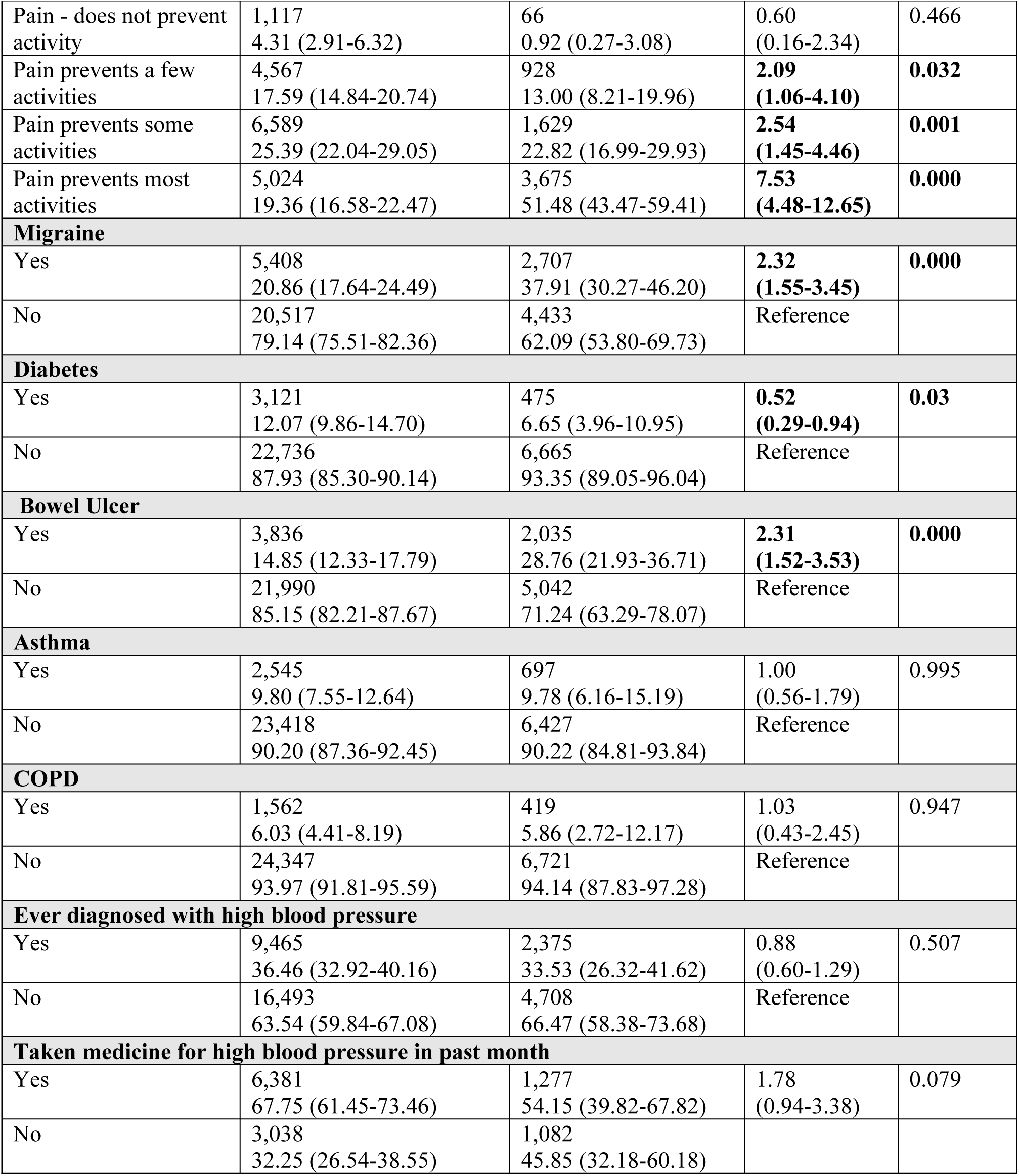
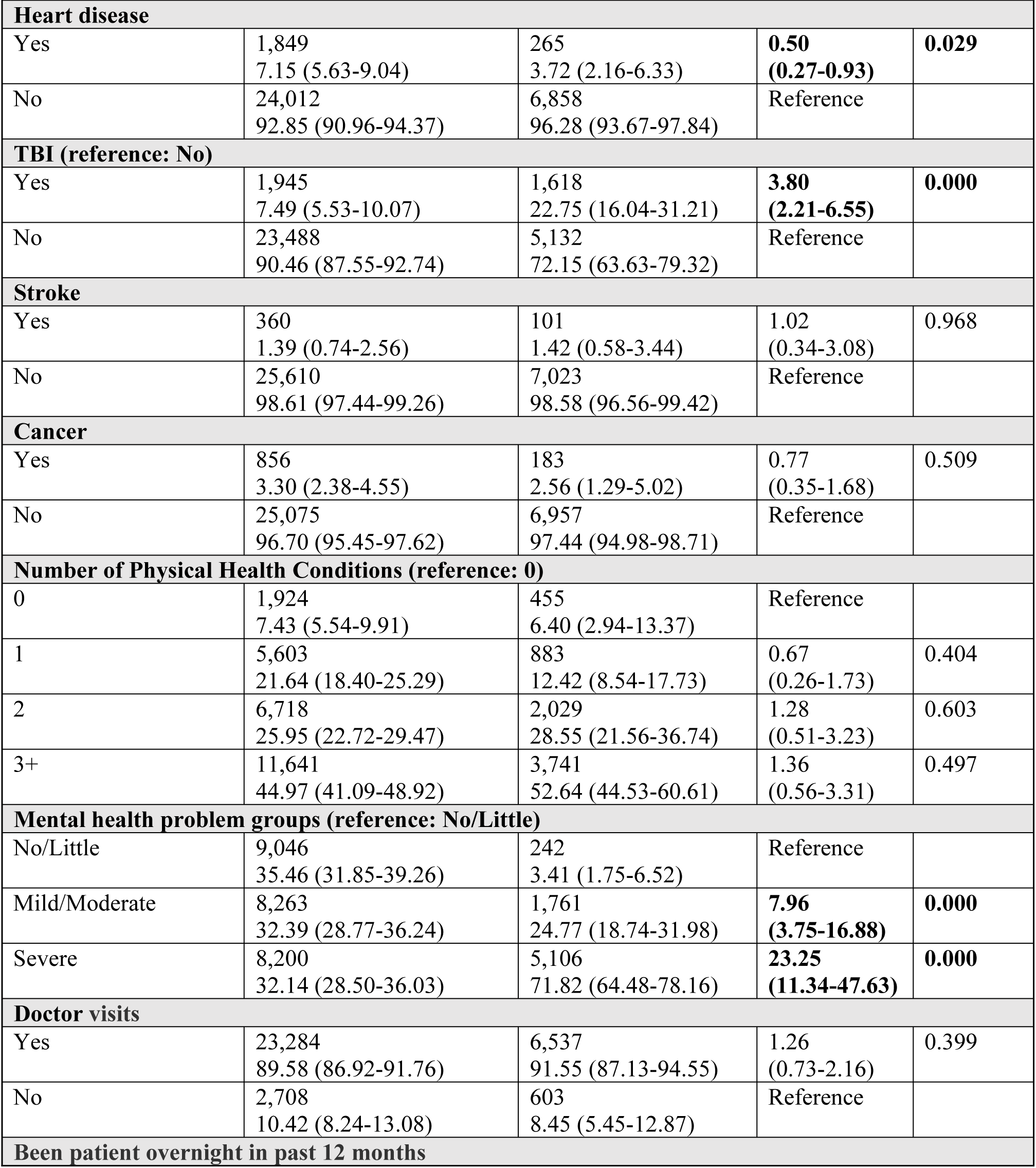
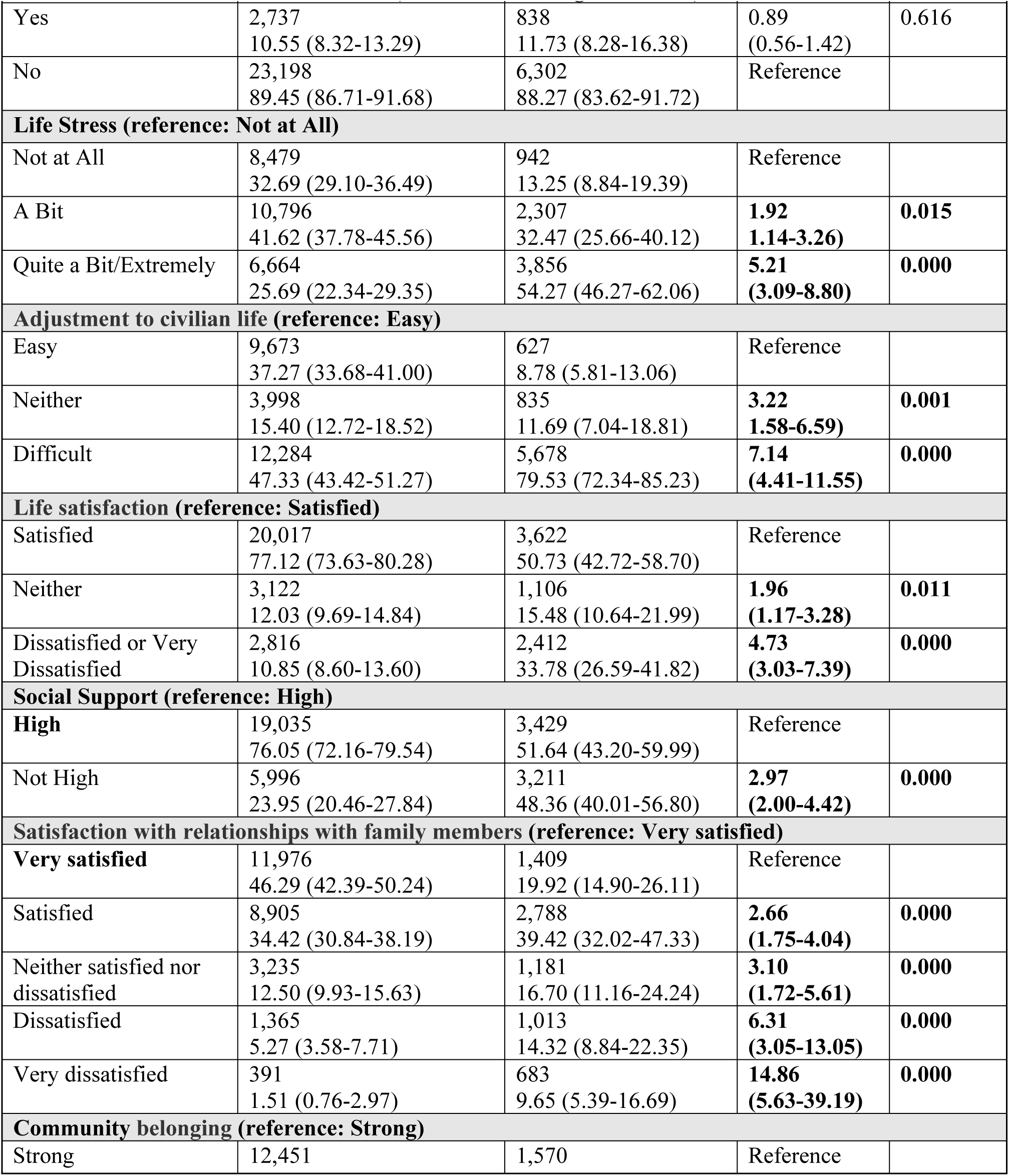
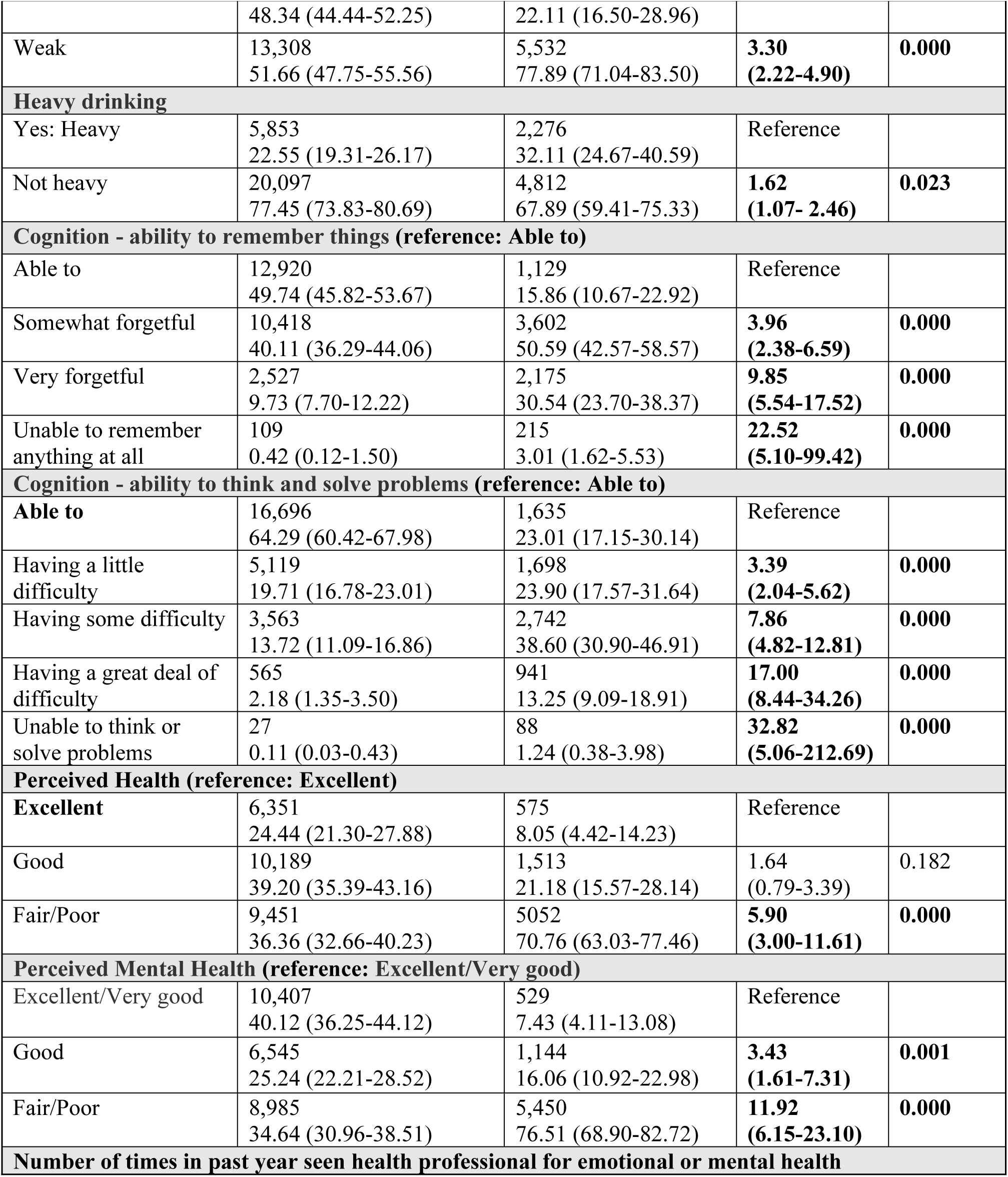
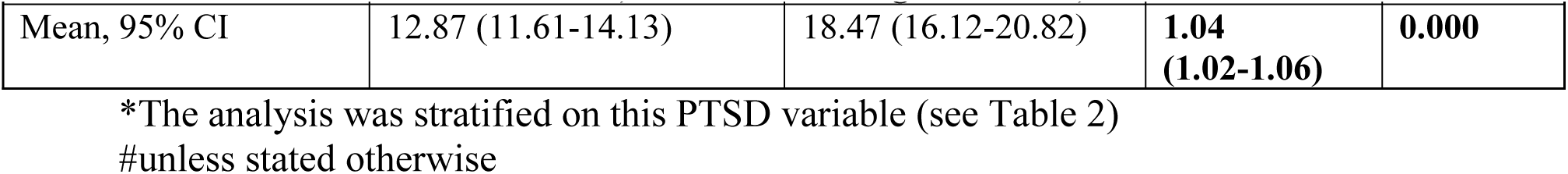
An unadjusted comparison of VAC clients with an active reimbursement for CMP (yes versus no) in 2019 using weighted estimates from the LASS 2019 survey.

### Demographic and Military Characteristics

The following variables were considered:

(i) Demographics, including age at the LASS 2019 survey, sex at birth, education status, marital status (married, living common-law, widowed, separated, divorced, and single, never married), province of residence, number of people who live in the household (including those 18 years old or younger), and total household income.
(ii) Service-related characteristics, including rank at release (Officer, Senior NCM, Junior NCM), excluding Entry Ranks – (Recruit, Officer Cadet, 2nd Lieutenant), service environment at release (Air [Air Force], Land [Army], Sea [Navy]), main activity (work/reserves, retired, disability, other), being ever deployed during military service, and length of service years of regular force service (calculated from the first date of enrolment to the last date of release. Length of service years was categorized into more than 20 years, less than 10 years and from 11 to 19 years [19].

#### Mental Health and Physical Health Conditions

Chronic conditions were assessed using the question: “Do you currently have any of the following diagnosed conditions (with more than 6 months of duration) by a health professional?” Response options included depression, anxiety, PTSD, hypertension, heart conditions, stroke, migraine, traumatic brain injury (TBI), Alzheimer’s disease, diabetes, gastrointestinal conditions, cancer, asthma or COPD, and hearing concerns [19]. Additional variables included pain, health status, having a regular medical doctor, being a patient overnight in the past 12 months, and the number of times in the past year being seen by a health professional for emotional/mental health.

### Psychosocial Factors and Cognitive Status

The following variables were considered: life stress, adjustment to civilian life, life satisfaction, social support, satisfaction with relationships with family members, community belonging, heavy drinking, Kessler Psychological Distress Scale (K10), and cognitive status (ability to remember things and ability to think and solve problems).

### Statistical Analysis

Descriptive statistics and bivariate modelling were used to assess unadjusted associations between various veteran characteristics and well-being indicators, and an active authorization for CMP as of March 2019.

For variable selection, a backward elimination process was used, including three blocks of independent variables: the first block included demographic and military factors; the second included life satisfaction, social support, seen or talked to a social worker and seen or talked to a psychologist; and the third included health status for specific comorbid conditions. For the total sample, to avoid multicollinearity, only one of the available PTSD variables, which was also used for stratification, was included in a backward elimination process. The statistical threshold used for the variable selection was defined as p ≤ 0.05.

Finally, using a multivariable logistic regression approach, we evaluated the association between the final set of variables and the authorization for CMP reimbursement for the total sample and by PTSD status, given this high prevalence and the strong bivariate association between PTSD and CMP reimbursement. The present work followed the same methods and definitions reported previously for this type of analysis [2]. The interaction between the PTSD status and sex was considered *a priori*, but was reported in the final model as not significant (p=0.787).

### Weighted Estimates

The study design includes the application of survey weights to generate findings that are representative of the target Veteran population. This survey weight accounts for unequal probabilities of selection, eligibility, and non-response. In all cases, weighted estimates using Taylor linearization were produced to account for the complex sampling design of the LASS 2019 survey in accordance with previously established methodology[19,21].

## Results

Of a total of 1,289 (a weighted estimation of 33,131) survey responders (87.6% males and 21.5% 60 years and older), the frequency of clients authorized to be reimbursed for CMP as of March 31, 2019, was 18.4% (n=238), corresponding to a weighted estimation of 7,140 clients. Table 1 presents the unadjusted associations between CMP 2019 authorizations and the variables of interest per domain.

An important aspect to highlight from the crude associations is that the self-reported PTSD diagnosis and the PTSD screener questions (responded positivity) were highly prevalent (65% to 82%) and associated with about three to six times higher odds of being authorized for CMP compared to those who did not report the symptoms or diagnosis (OR for self-reported PTSD diagnosis of 6.17; CI 95%: 4.21-9.03) (see Table 1). Given this high prevalence and the strong bivariate association, the multivariate analysis was performed for the total sample and using the subpopulation estimation method by PTSD status [20]. Of note, in unadjusted analyses, anxiety (OR of 5.91; 95% CI: 4.06-8.60) and mood (OR of 5.81; 95% CI: 3.89-8.68) disorders were also associated with being authorized for CMP, with a progressive increase in the strength of associations with CMP as the severity of mental health problems increased (see Table 1). In terms of physical health, traumatic brain injury (OR of 3.80; 95% CI: 2.21-6.55), migraines (OR of 2.32; 95% CI: 1.55-3.45), and bowel ulcers (OR of 2.31; 95% CI: 1.52-3.53) were more likely to be authorized for CMP, whereas, those with diabetes (OR of 0.52; 95% CI: 0.29-0.94) or heart diseases (OR of 0.50; 95% CI: 0.27-0.93) were less likely. Additionally, odds ratios were greater in New Brunswick (OR of 3.41; 95% CI: 1.90-6.10) and smaller in Quebec (OR of 0.30; 95% CI: 0.15-0.58) compared to Ontario. Different predictors were retained in the final logistic models. Table 2 presents the results of the final multivariate models for the total sample and each of the PTSD categories.

**Table 2.** Multivariable logistic regression models for the total sample and by the PTSD status*. Estimates are presented as odds ratios (OR) and 95% confidence intervals (CI).

| Variables | Total |  |  | No PTSD |  |  | PTSD |  |  |
| --- | --- | --- | --- | --- | --- | --- | --- | --- | --- |
|  | OR | 95% CI | P-value | OR | 95% CI | P-value | OR | 95% CI | P-value |
| Age in 2019, years (reference: ≥60) |  |  |  |  |  |  |  |  |  |
| <40 | 4.78 | 2.24-10.21 | 0.000 | 31.04 | 2.60- 369.90 | 0.007 |  |  |  |
| 40-49 | 3.23 | 1.62-6.44 | 0.001 | 11.62 | 1.26- 106.98 | 0.030 |  |  |  |
| 50-59 | 1.72 | 0.97-3.08 | 0.066 | 6.84 | 0.80- 58.85 | 0.080 |  |  |  |
| Sex# |  |  |  |  |  |  |  |  |  |
| Female vs Male | 0.79 | 0.36-1.73 | 0.562 |  |  |  |  |  |  |
| Environment of Service (reference: Air) |  |  |  |  |  |  |  |  |  |
| Land | 2.07 | 1.22-3.50 | 0.007 |  |  |  |  |  |  |
| Sea | 1.22 | 0.61-2.47 | 0.573 |  |  |  |  |  |  |
| Length of Service (reference: > 20 Years) |  |  |  |  |  |  |  |  |  |
| <10 |  |  |  |  |  |  | 4.46 | 1.89-10.51 | 0.001 |
| 10-19 |  |  |  |  |  |  | 3.73 | 2.10-6.63 | 0.000 |
| Labour Status (reference: Employed) |  |  |  |  |  |  |  |  |  |
| Unemployed | 1.02 | 0.32-3.27 | 0.967 | 0.04 | 0.002-0.910 | 0.042 | 1.30 | 0.24-7.07 | 0.759 |
| No job, not looking (and unable to work) | 3.10 | 1.78-5.40 | 0.000 | 5.42 | 1.41-20.80 | 0.014 | 2.41 | 1.19-4.88 | 0.015 |
| Posttraumatic stress disorder |  |  |  |  |  |  |  |  |  |
| Yes vs No | 2.81 | 1.69-4.66 | 0.000 |  |  |  |  |  |  |
| Anxiety |  |  |  |  |  |  |  |  |  |
| Yes vs No | 2.32 | 1.45-3.70 | 0.000 |  |  |  |  |  |  |
| Witness destruction during military service |  |  |  |  |  |  |  |  |  |
| Yes vs No |  |  |  |  |  |  | 2.52 | 1.21-5.24 | 0.014 |
| Suicidal Ideation in the past year |  |  |  |  |  |  |  |  |  |
| No vs. Yes |  |  |  |  |  |  | 2.21 | 1.24-3.94 | 0.007 |
| Pain Health Status (reference: No pain or discomfort) |  |  |  |  |  |  |  |  |  |
|  | OR | 95% CI | P-value | OR | 95% CI | P-value | OR | 95% CI | P-value |
| Pain does not prevent activity | 0.39 | 0.10-1.55 | 0.180 | 0.32 | 0.03-3.27 | 0.334 | 0.60 | 0.11-3.33 | 0.563 |
| Pain prevents a few activities | 2.04 | 0.94-4.44 | 0.071 | 13.42 | 1.37-131.70 | 0.026 | 2.33 | 0.86-6.33 | 0.096 |
| Pain prevents some activities | 1.87 | 0.97-3.62 | 0.062 | 6.14 | 0.93-40.43 | 0.059 | 1.70 | 0.66-4.38 | 0.275 |
| Pain prevents most activities | 3.61 | 1.97-6.60 | 0.000 | 21.33 | 2.97-153.30 | 0.002 | 3.01 | 1.34-6.77 | 0.008 |
| Number of Physical Health Conditions (reference: 0) |  |  |  |  |  |  |  |  |  |
| 1 |  |  |  | 1.27 | 0.06-26.65 | 0.876 |  |  |  |
| 2 |  |  |  | 17.25 | 1.14-260.11 | 0.040 |  |  |  |
| 3+ |  |  |  | 3.66 | 0.21-63.39 | 0.372 |  |  |  |
| Life Stress (reference: Not at All) |  |  |  |  |  |  |  |  |  |
| A Bit |  |  |  | 0.11 | 0.01-0.83 | 0.033 |  |  |  |
| Quite a Bit/Extremely |  |  |  | 0.09 | 0.01-0.80 | 0.031 |  |  |  |
| CMH_010: No. times in past year - seen health prof. emotional/mental health |  |  |  | 1.08 | 1.02-1.14 | 0.009 |  |  |  |

\*This model was additionally adjusted for the province of residence.
#For the total sample, interaction term between PTSD and sex was considered in the final model but was not statistically significant (p=0.787).

For the total sample, compared to those aged ≥60, younger age groups (<40 and 40-49 years) were significantly more likely to be authorized for the CMP reimbursement, with the greater odds for those<40 (OR of 4.78; 95% CI: 2.24-10.21). Other variables that were significantly associated with higher odds of authorization for CMP reimbursement were serving in a land environment (OR versus air of 2.07, 95% CI: 1.22-3.50), reporting "no job, not looking (and unable to work)" (a category that was distinct from being "unemployed"; OR versus employed of 3.10, 95% CI: 1.78-5.40), having PTSD (OR of 2.81, 95% CI: 1.69-4.66) and anxiety (OR of 2.32, 95% CI: 1.45-3.70) as well as pain, which prevents most activities (OR versus "no pain or discomfort" of 3.61, 95% CI: 1.97-6.60).

In stratified analyses, for those without PTSD, variables retained in the final model that were significantly associated with authorization for CMP reimbursement were age below 49 years old (versus 60 years and older), reporting "no job, not looking (and unable to work)" (versus employed) and pain, which prevents most activities (versus "no pain or discomfort") as well as suffering from two physical health conditions, seeing more frequently health professionals for emotional/mental health, but at the same time reported no life stress.

On the other hand, for those with PTSD, having spent 20 or fewer years in service (versus > 20 years), reporting "no job, not looking (and unable to work)" (versus employed), pain which prevents most activities (versus "no pain or discomfort"), witness destruction during military service and suicidal ideation in the past year (versus not) were retained in the final model and significantly associated with CMP reimbursement.

Notably, reporting "no job, not looking (and unable to work)" (versus employed) and pain, which prevents most activities, were retained in the total sample and both subgroups by PTSD status.

## Discussion

In this large cross-sectional study, linking survey and administrative data on a representative sample of Canadian veterans released from the Canadian Regular Force (1998-2018), several sociodemographic, health, and service-related factors were associated with authorization for CMP reimbursement.

We found that about one-fifth of veterans were authorized for CMP reimbursement, with particularly strong associations observed for variables related to PTSD, anxiety and mood disorders (unadjusted ORs>5) and severe pain (OR>7.5). From a physical health standpoint, individuals reporting traumatic brain injury, migraines and bowel ulcers were more likely to be authorized for CMP, whereas those with diabetes or heart diseases were less likely. Considering that these last two conditions have been associated with certain risks for complications following regular cannabis use [22,23], results from the current analyses could possibly point to contraindications preventing healthcare providers from authorizing cannabis for people with this medical profile. Unadjusted analyses also unveiled associations with poorer relations and social supports, as well as provincial differences, with significantly lower CMP authorizations in Quebec, which has a global regulatory approach known to be especially conservative, compared to Ontario. How different levels of the social environment, from individual relationships to political views about cannabis, may influence medical use should be further investigated under psychosocial and health equity lenses. Although retired individuals were twice as likely to be authorized for CMP than those actively working or on reserve, being on disability leave was the work status that was the most strongly associated with CMP (OR>6).

In multivariate analysis, younger age, inability to work, disabling pain that prevents most activities, PTSD and anxiety emerged as key correlates of CMP authorization in the overall sample.

Similar to what was seen in unadjusted analyses, inability to work and severe disabling pain were consistent correlates across PTSD strata, but additional associations differed by PTSD status. Among veterans without PTSD, younger age, multiple physical conditions, and frequent mental health service use were notable correlates, whereas among those with PTSD, fewer years of service, witnessing destruction during service, and suicidal ideation were associated with CMP authorization. These findings suggest that CMP authorization patterns may reflect not only mental and physical health conditions, but also broader functional limitations and service-related exposures.

Our results are consistent with prior studies documenting that younger veterans are more likely to use cannabis compared to older veterans [24]. However, while younger veterans (e.g., ages 18–25) have previously been shown to have higher overall rates of cannabis use, older veterans (especially those aged 60 and above) were more likely to report using cannabis specifically for medical reasons, such as pain management and mental health conditions [25]. The strong associations with inability to work and pain are also in line with previous reports linking cannabis use to self-management of chronic pain [26] and functional impairments [27]. The observed associations with PTSD and anxiety also mirror findings from other studies [15–18]. In veteran populations, cannabis use has been described as a strategy for coping with intrusive symptoms and hyperarousal [28]. However, cannabis use in the context of mental health disorders remains controversial, with some evidence suggesting potential short-term symptom relief but limited evidence of long-term efficacy and growing concerns regarding dependence, psychiatric comorbidity, and cognitive effects [1,29,30]. Longitudinal studies are urgently needed to assess longer-term effects and determine whether evidence supports these concerns.

The stratified analyses by PTSD status highlight the complex interplay of mental health, service experiences, and functional status. In Veterans without PTSD, CMP authorization was more strongly associated with markers of poor physical health and healthcare engagement, whereas in those with PTSD, service-related trauma and suicidality were key correlates. This distinction may reflect differing pathways to CMP authorization: for some Veterans, cannabis may be pursued primarily for physical health conditions, while for others, it may be linked to trauma and its sequelae. However, pain and functional disability seem to be key factors, regardless of the physical or psychological nature of health issues.

A key strength of this study is the use of large, population-level administrative data linked to the survey, which minimizes recall bias and captures real-world patterns of CMP authorization across a representative veteran population in Canada. The high response (72%) and share (92%) rates minimize selection bias. The inclusion of a broad range of variables, including physical and mental health conditions, functional limitations, and service-related exposures, provides a more comprehensive view than many prior studies, which have typically focused on narrower domains. The stratification by PTSD status represents a novel contribution, uncovering differences in correlates that were obscured in pooled analyses. Furthermore, this study examined CMP authorization rather than self-reported cannabis use, thereby offering unique insights into the regulated, physician-mediated process of access to medical cannabis. Together, these strengths enhance the policy and clinical relevance of our findings, particularly in the Canadian context, where authorization is tied to reimbursement through Veterans Affairs Canada.

Several limitations should be acknowledged. First, the cross-sectional design precludes causal inference; associations cannot establish whether these factors drive CMP authorization or instead reflect the characteristics of those already authorized. Second, reliance on administrative data and survey may underrepresent symptom burden, functional impairment, or informal cannabis use. Third, although we included a broad set of potential correlates, residual confounding remains a possibility. For example, reliance on self-reported data, including PTSD diagnosis and symptoms, pain severity, and employment status, introduces potential for recall and social desirability bias and lacks clinical validation. Importantly, information on actual cannabis use and substance use problem/disorder was not available. Nevertheless, the large sample size and use of data sources based on reimbursement strengthen the robustness of our findings.

Building on these study findings, future studies should prioritize longitudinal designs to establish relationships between potential predictors and CMP authorization/cannabis use, as well as to evaluate CMP’s real-world effectiveness on outcomes highly relevant for veteran (e.g., pain interference, PTSD symptoms, functional recovery) and identify individuals who may benefit the most. For policy makers, distinct correlates by PTSD status highlight the importance of integrated pain-mental health services and careful benefit-risk discussions regarding cannabinoids, particularly where poly-morbidity and functional limitations are present.

## Conclusion

In this large cross-sectional study on a representative sample of Canadian veterans, CMP authorization among Canadian veterans was most strongly associated with younger age, inability to work, severe disabling pain, and mental health conditions, with notable differences between those with and without PTSD. These results underscore the need for careful assessment of functional status, service exposures, and mental health in evaluating cannabis authorization. Longitudinal studies are needed to determine how these factors influence long-term outcomes and to guide evidence-based policies for CMP prescribing and reimbursement.

## Data Availability

The data that support the findings of this study were obtained from Veterans Affairs Canada (VAC). Due to ethical, legal and privacy restrictions, these data are not publicly available. De-identified aggregate data and analysis code may be available from the corresponding author on reasonable request.

## Supporting Information

**Table S1. List of variables considered for inclusion in the statistical model.**

**Figure S1. Administrative cannabis for medical purposes (CMP) authorizations database as of March 31, 2019, and the Life After Service Survey 2019 data linkage.**

